# Menstrual health, food insecurity, and protection vulnerability among adolescent girls in internally displaced persons camps in Benue State, Nigeria: A focused ethnographic study

**DOI:** 10.64898/2026.10.07.26365008

**Authors:** Ogechi Cecilia Ofor, Felicity Roux, Samson Abanni, Ogodo Lucy Uzodimma, Chinenye Magdaline Obiorah, Mimidoo Mirabelle Uhundu, Ogbu Chidimma, Muizat Bukola Olatunde, Cynthia Uloma Offor, Fahis Taiwo Akoshile, Olaide Zainab, Ofor Chiamaka Marycynthia, Ofor Ngozi Agatha, Patrick Maduabuchi Aja

## Abstract

Adolescent girls living in internally displaced persons (IDP) camps manage menstruation amid food insecurity, inadequate water, sanitation and hygiene (WASH), disrupted education, gendered social norms and protection systems. These challenges are often addressed through separate humanitarian programmes, obscuring how they interact in girls’ everyday lives. This study explored menstrual-health experiences and perceived nutritional deprivation among adolescent girls in five IDP camps in Benue State, Nigeria. We conducted a focused ethnographic qualitative study in the International Market, Mega, Ortese, Uikpam, and Abagana camps. Twenty-five in-depth interviews with menstruating adolescent girls aged 13–19 years were complemented by focus group discussions with adolescent girls, adult women, adult men, and camp leaders, and structured WASH and food observations. Interviews and discussions were conducted in English, Tiv or both, transcribed verbatim and translated where necessary. Data were managed in MAXQDA and analysed using reflexive thematic analysis. Frequencies describe the distribution of 435 coded excerpts and are not participant prevalence estimates. Six themes and 20 subthemes were generated. Menstrual-health management and material constraints accounted for 95 coded excerpts (21.8%), followed by knowledge and lived experience (80; 18.4%), food insecurity and perceived nutritional deprivation (70; 16.1%), coping and adaptive agency (70; 16.1%), sociocultural beliefs and taboos (65; 14.9%), and self-reported symptoms and participation restrictions (55; 12.6%). Girls described irregular access to commercial pads, reliance on improvised materials, limited water and soap, and toilets that were unsafe or inaccessible at night. Dietary monotony, meal skipping, and hunger were perceived to intensify pain, weakness, and fatigue. Fear of staining, menstrual silence, and household restrictions contributed to withdrawal from school, livelihood, and domestic activities. Peer support was protective, but some girls undertook informal work to obtain menstrual materials, raising safeguarding concerns. Menstrual health in prolonged displacement is not an isolated hygiene issue but an outcome of interacting material, nutritional, sociocultural, educational, and protection constraints. Humanitarian responses should combine predictable access to acceptable menstrual materials with safe WASH facilities, adolescent-responsive food and nutrition support, confidential health services, community education and safeguarding mechanisms. Reported symptoms and nutritional experiences were not clinically measured.

## Introduction

Menstrual health is a state of physical, mental and social well-being in relation to the menstrual cycle, rather than simply the absence of menstrual problems. Achieving it requires accurate knowledge, access to preferred materials, water and sanitation, timely health care, freedom from stigma, and the ability to participate in education, work and social life [1]. A qualitative metasynthesis from low- and middle-income countries similarly showed that menstrual experiences are shaped by the interaction of sociocultural context, knowledge, material resources, facilities, interpersonal support and individual confidence [2].

These enabling conditions are frequently disrupted in humanitarian settings. Reviews describe shortages of acceptable materials, inadequate privacy, unsafe toilets, poor disposal systems, limited information and weak coordination across health, protection and WASH sectors [3–6]. Recent qualitative studies provide direct illustrations: adolescent girls in Palestinian refugee camps reported inadequate preparation, menstrual secrecy and divergent expectations among girls, mothers and teachers, while internally displaced girls in Tigray described pad shortages, poor latrines, lack of privacy and exchanges of menstrual materials for essential commodities [7,8]. Such constraints are particularly consequential for adolescents whose first menstrual experiences may occur during displacement and whose autonomy depends on caregivers, schools and humanitarian distributions.

Studies in refugee and low-resource settings show that menstrual health depends on the interaction of material access, knowledge, social support and usable facilities. In the Nakivale refugee settlement in Uganda, adolescent girls described material scarcity, pain, stigma and difficulties maintaining privacy and hygiene [9]. Research from Cox’s Bazar demonstrated that menstruation-supportive WASH requires safe location, lighting, locks, water, washing space and culturally acceptable disposal arrangements, not simply the presence of toilets [10]. A qualitative assessment across secondary schools in Uganda likewise found menstrual-health barriers at individual, interpersonal, institutional and societal levels [11]. Across Africa, a recent scoping review concluded that provision for displaced women and girls remains inconsistent and insufficiently integrated with other humanitarian services [5].

Internal displacement in Nigeria creates additional structural constraints. Reviews and primary studies identify weak access to reproductive-health information and services, gendered insecurity, overburdened humanitarian provision and limited opportunities for confidential adolescent care [12–16]. Recent work in north-central Nigeria documented financial, geographic and sociocultural barriers to sexual and reproductive health care and reliance on potentially harmful coping practices, while a 2025 qualitative study found continuing gaps in adolescent-responsive information, confidentiality and service delivery in an IDP camp [17,18]. Menstrual maintenance, laundering and disposal challenges have also been documented among displaced girls and women in north-eastern Nigeria [19]. However, evidence remains geographically concentrated and often treats menstrual health, food insecurity, WASH and protection as separate domains.

Nutrition is especially important during adolescence, when growth and menstrual blood loss increase nutritional demands. A systematic review found that adolescent girls in low- and middle-income countries commonly experience monotonous diets and inadequate micronutrient intake [20]. Iron deficiency and anaemia can contribute to fatigue and impaired functioning, although diagnosis requires appropriate clinical or laboratory assessment [21]. In displacement settings, food insecurity may therefore shape how girls experience and interpret menstrual pain, weakness and participation, but qualitative perceptions should not be equated with measured malnutrition or anaemia.

Nigeria-focused evidence shows persistent gaps in menstrual knowledge, access to materials, privacy, supportive communication, and policy coordination [22,23]. Less is known about how adolescent girls in Benue State IDP camps understand the combined effects of displacement, hunger, menstrual-material scarcity, WASH conditions, sociocultural norms, and protection risks. This study therefore explored menstrual-health experiences among adolescent girls in five Benue State IDP camps. It examined how food insecurity, environmental conditions, community beliefs, and coping practices shaped menstrual well-being, dignity, and participation.

## Materials and methods

### Ethics Statement

Ethical approval was obtained from the Benue State Emergency Management Agency Ethical Review Board (approval number: MOH/STAT/2204/1/205/2026) on 2 March 2026. Permission to enter each camp was additionally granted by the Benue State Emergency Management Agency (SEMA) and camp leadership. For participants aged 13–17 years, written informed consent was obtained from a parent or legal guardian, and assent was obtained from the adolescent. Participants aged 18–19 years provided their own written informed consent. Participation was voluntary, withdrawal was permitted without penalty, and interviews were conducted privately. Identifiers used in reporting combine data-source and camp codes and do not disclose participant names.

### Study design and setting

A focused ethnographic qualitative design was used to examine everyday menstrual-health experiences within the social and material environment of prolonged displacement. The study was conducted in five purposively selected IDP camps in Benue State, Nigeria: International Market, Mega, Ortese, Uikpam and Abagana. The sites represented different camp sizes, histories, and service environments and had active adolescent populations. Fieldwork was undertaken from April 2026 to June 2026 following ethical approval obtained on 2 March 2026.

### Participants and recruitment

Participant recruitment was conducted from 18 November 2025 to 10 March 2026. Participants were selected purposively because they had direct experience relevant to the research questions. Adolescent participants were eligible when they were aged 13–19 years, had experienced menstruation, and had lived in the camp for at least three months. Camp leaders, women leaders, and community mobilisers assisted the study team in identifying potentially eligible participants; researchers then independently explained the study and obtained consent or assent. Twenty-five adolescent girls completed in-depth interviews (IDIs). Focus group discussions (FGDs) involved adolescent girls, adult women, adult men, and camp leaders. Recruitment continued until the research team judged that the dataset provided adequate depth and variation across camps and participant groups. The study involved five internally displaced persons (IDP) camps. Four girls were selected from each camp, giving a total of 20 girls. For the focus group discussions (FGDs), 25 girls participated. They were organized into five focus groups, with one group drawn from each of the five IDP camps and approximately five girls participating in each group.

### Data collection

Three complementary methods were used: semi-structured IDIs, FGDs, and structured participant observation. IDIs explored menstrual knowledge, menarche, material access, hygiene practices, food access, perceived nutritional challenges, symptoms, sociocultural beliefs, education and livelihood participation, coping, and experiences of humanitarian services. Separate FGD guides were used for adolescent girls, adult women, adult men, and camp leaders to examine community norms, household practices, institutional responses, and differences in perspective. Interviews and discussions lasted approximately 60 minutes and were conducted in private locations within the camps.

Observation checklists documented water points, toilets, bathing spaces, availability of soap and lighting, privacy, menstrual waste disposal, food availability, and other environmental conditions relevant to menstruation. With permission, IDIs and FGDs were audio-recorded. Researchers maintained field notes on non-verbal communication, interactions, and contextual observations during and immediately after data collection.

### Transcription and translation

Data collection was conducted in English, Tiv or a combination of both according to participant preference. Recordings in Tiv were transcribed in Tiv and translated into English by bilingual research assistants. Selected transcripts underwent independent back-translation, and discrepancies were discussed by the research team. Culturally specific expressions were retained where necessary to preserve meaning.

### Data analysis

Data were managed in MAXQDA and analysed using the six recursive phases of reflexive thematic analysis: familiarisation, coding, initial theme development, theme review, theme refinement and reporting [24]. Researchers read IDI transcripts, FGD transcripts, field notes and observation records repeatedly and coded meaningful segments line by line. Codes were compared across participant groups, camps and data sources and developed into candidate themes. The research team reviewed theme boundaries, divergent accounts and the relationship between experiential, sociocultural and structural findings.

The final analytic framework comprised 435 coded excerpts, 20 subthemes and six overarching themes. Code frequencies were used descriptively to show how coded material was distributed of illustrative quotations. Transferability was supported by a description of the setting and participant-selection procedures. Dependability and confirmability were strengthened through an audit trail, reflexive field notes and team discussion. The research team comprised Nigerian researchers with public health and community research experience. Familiarity with the local context supported rapport but required continuous reflection on assumptions, authority and the interpretation of culturally embedded accounts. Reporting was informed by the Consolidated Criteria for Reporting Qualitative Research [25].

## Results

### Overview of the thematic findings

The analysis generated six themes and 20 subthemes from 435 coded excerpts (Table 1). The largest share of coded material concerned menstrual-health management and material constraints (95 excerpts; 21.8%). Knowledge and lived experience contributed 80 excerpts (18.4%). Food insecurity and perceived nutritional deprivation, and coping and adaptive agency, each contributed 70 excerpts (16.1%). Sociocultural beliefs and taboos contributed 65 excerpts (14.9%), while self-reported symptoms and restrictions on participation contributed 55 excerpts (12.6%). These percentages describe excerpts rather than people.

**Table 1.** Themes generated from the focused ethnographic analysis.

| Theme | Constituent subthemes | Coded excerpts, n (%) | Data sources | Camp coverage |
| --- | --- | --- | --- | --- |
| Knowledge, perceptions and lived experience of menstruation | Pre-displacement support and menarche knowledge; emotional transformation in camp; disrupted normalcy, education and aspirations | 80 (18.4) | IDI, FGDs | All five camps |
| Menstrual-health management and material constraints | Inadequate materials; substandard WASH; nocturnal safety barriers; improvised hygiene practices | 95 (21.8) | IDI, FGDs, observations | All five camps; night-safety findings concentrated in International Market and Ortese |
| Food insecurity, perceived nutritional deprivation and menstrual well-being | Dietary monotony; perceived worsening with hunger; appetite disruption, meal skipping and food beliefs | 70 (16.1) | IDI, FGDs, observations | All five camps |
| Sociocultural beliefs, taboos and symbolic meanings | Silence and shame; household restrictions; spiritual beliefs about menstrual blood and disposal | 65 (14.9) | IDI, FGDs, observations | All five camps; ritual-disposal accounts most explicit in Abagana |
| Self-reported symptoms and participation in daily life | Pain, weakness, fatigue and self-described irritation/infection; restrictions on domestic and livelihood activities; withdrawal | 55 (12.6) | IDI, FGDs | All five camps |
| Coping, peer support and adaptive agency | Individual improvisation; peer solidarity; labour to obtain materials; ambivalence toward formal services | 70 (16.1) | IDI, FGDs, observations | All five camps; labour exchange reported in Uikpam and Abagana |
| Total | Six themes and 20 subthemes | 435 (100.0) | IDI, FGDs, observations | Five camps |

Note. Percentages are based on 435 coded excerpts, not participants. Code frequencies describe the distribution of the analysed text and are not prevalence estimates or measures of thematic importance. IDI = in-depth interview; FGD = focus group discussion; WASH = water, sanitation and hygiene.

### Theme 1: Displacement transformed menstruation from a manageable event into persistent uncertainty

Before displacement, participants commonly associated menstruation with maternal guidance, access to pads, private washing spaces and regular school attendance. One participant summarised this earlier experience as: “I had privacy, I had dignity, and I was hopeful about my future” (IDI-IM1). For girls who reached menarche after displacement, camp conditions constituted their first and only context for learning menstrual management. IDI-ME3 explained, “I started seeing my period in camp here. Back home, I had not started seeing my period.”

Across camps, the onset of menstruation was described as triggering an immediate calculation about materials and concealment. “When my period starts, the first thing I feel is worry. I start thinking about whether I have anything to use,” said IDI-IM1. Displacement also disrupted schooling, income-generating activities and family support. Participants linked these losses to reduced access to menstrual information, peer support and future opportunities.

### Theme 2: Material scarcity and unsafe WASH conditions undermined dignified menstrual management

Commercial pads were available irregularly, usually through episodic distribution or personal purchase. Participants described using folded cloth, tissue, cotton wool, hot-water-soaked rags and reusable pads. IDI-ME2 stated, “I don’t always have pads available, and there is no proper place to manage myself.” Knowledge and practices differed between camps, partly reflecting variation in NGO sensitisation. Some girls described sun-drying reusable cloth or pads, while others buried disposable materials because bins were unavailable.

Observations across the camps identified limited soap, distant water points, inadequate menstrual waste systems, and insufficient privacy. Lighting was absent around toilets in several sites, and some facilities were locked or inaccessible at night. At Ortese, one participant reported bathing in the open while clothed because toilets were locked. These environmental constraints made menstrual management time-dependent and sometimes pushed girls outside protected camp spaces.

### Theme 3: Food insecurity was perceived to intensify menstrual pain, weakness and fatigue

Participants in all five camps described diets dominated by a limited range of starchy staples. Market observations suggested that vegetables, beans, meat and other nutrient-dense foods could sometimes be found near the camps but were frequently unaffordable. Girls and caregivers described meal skipping, reduced appetite during menstruation and household food allocation that prioritised younger children. A young mother explained, “Sometimes it is just small food, and I have to think about my child first before myself” (IDI-IM4).

Participants consistently interpreted hunger as worsening menstrual discomfort and reducing their ability to function. IDI-OR3 said, “When food is not enough, I feel weaker, and the pain feels worse.” A women’s FGD participant similarly observed that a hungry menstruating girl became weak and unable to function, “but we overlook [it] because food is limited.” These accounts establish perceived links between food insecurity and menstrual well-being, not clinical diagnoses of malnutrition, iron deficiency or anaemia.

### Theme 4: Menstrual silence and taboos restricted disclosure, while education created gradual change

Menstruation was widely described as private and difficult to discuss. Girls feared ridicule, visible staining and being judged as unclean. Men’s and women’s FGDs described household restrictions in some families, including prohibitions on cooking, sharing sleeping spaces or interacting with culturally designated men during menstruation. At Abagana, girls described beliefs that menstrual blood could be used for harmful spiritual purposes, encouraging hidden disposal practices.

Norms were not uniform or static. Participants in camps with sustained NGO education reported greater communication between mothers and daughters and increased willingness among girls to ask questions. A women’s FGD participant at Mega explained that menstruation had previously been difficult to discuss, but awareness programmes were helping mothers change how they communicated with their daughters. Participants attributed this gradual change to repeated awareness activities and increased communication between mothers and daughters.

### Theme 5: Menstrual difficulties constrained daily participation and psychosocial well-being

Participants reported abdominal pain, weakness, tiredness, irritation and experiences they described as “infection.” Because no clinical examination or laboratory testing was performed, these accounts are reported as self-described symptoms rather than confirmed diagnoses. Participants perceived more severe difficulties when materials, water and privacy were limited. Several girls described being unable to fetch water, collect firewood, farm, complete domestic work or attend community activities during menstruation.

Fear of staining contributed to staying indoors, becoming quiet and avoiding social contact. One girl explained, “I feel scared moving about because I might sit somewhere and get stained” (FGD-AG-R5). In a camp economy where girls’ labour contributed to household access to water, food and fuel, temporary inability to participate affected more than the individual.

### Theme 6: Girls exercised agency, but some coping strategies generated new protection risks

Girls described rest, frequent bathing, cloth use, improvised materials and advice from health teams as ways of managing limited resources. Peer solidarity was especially visible in Abagana and Mega, where girls shared pads, information and “older-sister” support. One participant said she asked “a friend that won’t laugh at me” when she needed a pad. These networks reduced isolation but could not compensate for recurrent material shortages.

At Uikpam and Abagana, participants described casual labour undertaken to buy pads or meet basic needs. A women’s FGD participant stated, “Some girls go out to work for people just to get money for pads. It is risky, but they don’t have a choice.” The data did not establish transactional sex or exploitation in exchange for menstrual materials; however, unsupervised labour outside camp environments created plausible safeguarding concerns, particularly in a setting where women leaders also recounted violence during livelihood activities.

Formal services were viewed ambivalently. Participants valued NGO distributions and education but described services as irregular and overwhelmed. “Many people, not enough attention” was a recurring characterisation. Some girls became reluctant to disclose menstrual needs because they expected little response. These findings suggest that predictable, confidential and adolescent-responsive support may matter as much as one-off distributions.

## Discussion

### Principal findings

This study shows that menstrual health among adolescent girls in Benue State IDP camps was produced through an interlocking system of displacement-related deprivation. Material scarcity, unsafe WASH conditions, food insecurity, menstrual stigma, educational disruption, and limited services did not operate independently; together, they shaped worry, concealment, self-reported symptoms, participation, and coping. These findings converge with recent evidence from internally displaced adolescent girls in Ethiopia and refugee adolescents in Palestine, while extending the literature by showing how hunger, menstrual-material poverty and informal labour were experienced within the same everyday system [7,8].

### Material and WASH constraints

The findings reinforce evidence that product provision alone cannot achieve menstrual health in humanitarian settings. Girls require acceptable materials, water, soap, privacy, secure doors, lighting, washing and drying options, and culturally acceptable disposal systems [5,10,19,26].

Similar shortages of materials, privacy and functional latrines were recently reported among internally displaced adolescent girls in Tigray [8]. When toilets were distant, locked or unsafe at night, theoretical access did not translate into practical access. Humanitarian WASH indicators should therefore assess usability, safety and privacy throughout the day and menstrual cycle rather than simply count facilities.

### Food insecurity and perceived nutritional deprivation

Participants’ accounts extend menstrual-health research by locating menstrual pain and fatigue within the daily realities of insufficient and monotonous food. Existing evidence demonstrates poor dietary quality among many adolescent girls in low- and middle-income settings and identifies iron status as relevant to fatigue and cognitive or physical functioning [20,21]. However, the present study did not measure dietary intake, anthropometry, haemoglobin or biomarkers. The findings should therefore be interpreted as lived and socially shared explanations of bodily experience. Future studies should integrate qualitative accounts with validated food-security measures, dietary assessment and clinically appropriate screening.

### Stigma, education and adaptive agency

Menstrual silence restricted help-seeking and reinforced anxiety about staining and disclosure. At the same time, girls and caregivers described norms changing where community education was sustained. This aligns with studies in refugee camps and Ugandan schools showing that preparedness, confidence and participation depend on timely information, supportive communication and responsive institutions as well as products and facilities [2,7,9,11]. Programmes should engage girls, mothers, men, camp leaders and schools without treating local culture as fixed or uniformly harmful. Peer networks already operating within camps can be strengthened as sources of information and referral, while institutional responsibility must remain with service providers.

### Protection and service implications

The need to perform informal labour to obtain pads is an important protection finding. Period poverty becomes a safeguarding issue when access to an essential material depends on mobility, income or negotiation in environments with weak oversight. In Tigray, displaced adolescent girls described exchanging menstrual pads for life-saving commodities, and studies from Nigeria have documented transactional sex, gender-based violence, economic precarity and service barriers in displacement settings [8,16,18,27]. The present data did not demonstrate transactional sex in exchange for menstrual products; rather, they identify a credible pathway of vulnerability requiring prevention. A minimum integrated response should include predictable access to a choice of menstrual materials; safe, lit and lockable WASH facilities; confidential adolescent health support; food-security and nutrition assessment; referral for severe symptoms; protection monitoring; and accessible feedback mechanisms.

Coordination is essential. Nigerian studies point to structural gaps, financial and geographic barriers, inconsistent availability of services and overreliance on humanitarian and non-governmental actors for reproductive-health care in displacement settings [13,15–18]. Menstrual-health indicators should be incorporated into camp health, WASH, nutrition, education and protection plans, with adolescent girls participating meaningfully in service design and monitoring. One-off pad distributions may provide temporary relief but are unlikely to restore dignity, trust or participation when water, food, safety and confidential care remain inadequate.

### Strengths and limitations

The study drew on five camps, several participant groups and three complementary data sources. Interviews in participants’ preferred languages, translation checks, field observation, reflexive notes and team review increased contextual depth and interpretive credibility. The inclusion of men, women and camp leaders enabled comparison between girls’ lived experiences and wider household or institutional narratives.

Several limitations should guide interpretation. Purposive recruitment and assistance from camp leaders may have favoured participants who were more visible or comfortable engaging with researchers. Sensitive topics were vulnerable to social-desirability and recall bias, and group settings may have discouraged dissenting accounts. Translation may have altered nuance despite review and back-translation. The study did not include clinical assessment, anthropometry, haemoglobin testing, quantitative dietary measurement or validated food-security scales; consequently, it cannot estimate malnutrition, anaemia, infection or causal relationships. Code frequencies describe excerpts rather than participants and should not be interpreted as prevalence. Camp-level variation and the absence of a non-displaced comparison group also limit transferability.

## Conclusions

Adolescent girls in five Benue State IDP camps experienced menstruation within a system of material scarcity, food insecurity, inadequate WASH, stigma, disrupted participation and fragile support. Girls displayed considerable agency through improvisation, peer solidarity and selective use of humanitarian services, but some coping strategies exposed them to additional risk. Menstrual health should therefore be treated as a cross-sector humanitarian priority rather than a narrow pad-distribution activity. Integrated, girl-centred responses should combine acceptable menstrual materials, safe WASH, food and nutrition support, education, confidential clinical referral and safeguarding. Further mixed-methods research should test these pathways using validated nutritional and health measures while preserving displaced girls’ accounts of dignity, agency and everyday survival.

## Data Availability

The qualitative data underlying this study contain sensitive information from adolescent girls and internally displaced persons and cannot be made publicly available because of ethical and confidentiality restrictions and the potential risk of participant identification. Relevant de-identified excerpts supporting the findings are included in the manuscript. Additional de-identified data or supporting analytic materials may be made available to qualified researchers upon reasonable request and subject to approval by the relevant institutional ethics committee and applicable participant confidentiality and data-protection requirements. Access requests should be directed to the appropriate institutional ethics or research oversight body responsible for the study

## Acknowledgments

The authors thank the adolescent girls, women, men, camp leaders, community mobilisers and humanitarian personnel who supported the study. The authors also acknowledge the Cecilia for Empowerment Foundation and Tomorrow is a Girl Initiative for their support of the research.

